# Simulation-based power analysis could improve the design of clinical trials in Alzheimer’s disease

**DOI:** 10.1101/2022.12.24.22283807

**Authors:** Daniel Andrews, Douglas L. Arnold, Danilo Bzdok, Simon Ducharme, Howard Chertkow, D. Louis Collins, the Alzheimer’s Disease Neuroimaging Initiative

## Abstract

Clinical trials of new treatments in different progressive diseases use power analysis to determine the sample size needed for a trial to obtain a statistically significant estimate for an anticipated treatment effect. In trials with parallel designs, the standard power analysis approach is based on a two-sample t-test. For example, the standard t-test approach was used in determining the sample size for the Phase 3 trials of aducanumab, the first drug approved by the United States Food and Drug Administration (FDA) to potentially slow cognitive decline in early-stage Alzheimer’s disease. However, t-tests contain normality assumptions, and t-test-based power analyses do not implicitly factor in the uncertainty about anticipated treatment effects that arises due to inter-subject heterogeneity in disease progression. These limitations may lead to recommended sample sizes that are too small, potentially making a trial blind to a treatment effect that is truly present if the cohort’s endpoints are not normally distributed and/or the anticipated treatment effect is overestimated.

To address these issues, we present a novel power analysis method that (1) simulates clinical trials in a progressive disease using real-world data, (2) accounts for inter-subject heterogeneity in disease progression, and (3) does not depend on normality assumptions. As a showcase example, we used our method to calculate power for a range of sample sizes and treatment effects in simulated trials similar to the Phase 3 aducanumab trials EMERGE and ENGAGE. As expected, our results show that power increases with number of subjects and treatment effect (here defined as the cohort-level percent reduction in the rate of cognitive decline in treated subjects vs. controls). However, inclusion of realistic inter-subject heterogeneity in cognitive decline trajectories leads to increased sample size recommendations compared to a standard t-test power analysis. These results suggest that the sample sizes recommended by the t-test power analyses in the EMERGE and ENGAGE Statistical Analysis Plans were possibly too small to ensure a high probability of detecting the anticipated treatment effect. Insufficient sample sizes could partly explain the statistically significant effect of aducanumab being detected only in EMERGE. We also used our method to analyze power in simulated trials similar the Phase 3 lecanemab trial Clarity AD. Our results suggest that Clarity AD was adequately powered, and that power may be influenced by a trial’s number of analysis visits and the characteristics of subgroups within a cohort.

By using our simulation-based power analysis approach, clinical trials of treatments in Alzheimer’s disease and potentially in other progressive diseases could obtain sample size recommendations that account for heterogeneity in disease progression and uncertainty in anticipated treatment effects. Our approach avoids the limitations of t-tests and thus could help ensure that clinical trials are more adequately powered to detect the treatment effects they seek to measure.

## 1. Introduction

### 1.1. Two-sample t-test power analysis in parallel-design clinical trials

Power analysis helps trial designers determine the sample size needed to obtain a statistically significant estimate for an anticipated treatment effect size at a given power and significance level.^1^ Randomized controlled clinical trials with parallel designs often use a two-sample t-test approach for power analysis. Such an approach was used in determining the sample size for the Phase 3 trials of aducanumab, the first drug approved by the United States Food and Drug Administration (FDA) to potentially slow cognitive decline in early-stage Alzheimer’s disease patients by attacking brain amyloid-beta plaques.^2,3^

For two-sample t-test power analyses, Cohen’s *d* is used to define the treatment effect size as the final observation standardized mean difference between treatment and control groups for a trial’s clinical or biological endpoint.^4^ Concretely, the effect size is defined as the anticipated final observation difference in endpoint means between treatment groups, divided by the anticipated pooled standard deviation for that endpoint. For example, if the anticipated difference in means (i.e., the treatment effect) is 0.5 and the pooled standard deviation is 1.92, then the treatment effect size (i.e., Cohen’s *d*) is 0.5/1.92 ≈ 0.26. The mean and standard deviation values are posited in reference to endpoint distributions measured in previous experiments or trial phases.

The significance level, α, is the probability of a Type 1 (false positive) error.^5^ In null hypothesis statistical testing, a Type 1 error has occurred when the null hypothesis is rejected when it is, in fact, true.^5^ For clinical trials, α can be conceptualized as the probability a trial will commit a Type 1 error by obtaining a statistically significant estimate for a treatment effect when one truly *is not* present.

Power is defined as 1 – β, where β is the probability of a Type 2 (false negative) error.^5^ In null hypothesis statistical testing, a Type 2 error has occurred when the null hypothesis is accepted but is actually false.^5^ For a clinical trial, power can be conceptualized as the probability the trial will obtain a statistically significant estimate for a treatment effect that truly *is* present. In clinical trial power analysis, power and significance levels are often quoted in units of percent. For an example trial with an anticipated treatment effect size of 0.26, a desired power of 0.9 (90%) at α = 0.05 (5%), and an overall 30% dropout anticipated, the sample size recommended by a two-sample, two-sided t-test power analysis is approximately 450 subjects per treatment group. We implemented this example in the widely used *R* statistical computing framework.^6,7^

### 1.2. The limitations of two-sample t-test power analysis for trial design in Alzheimer’s disease

Two-sample t-test power analysis is limited by its strong assumptions. A first assumption is that a trial’s endpoint will be normally distributed with equal variance in each treatment group. Like many other diseases, Alzheimer’s disease (AD) is an extremely heterogeneous disorder. AD has multiple possible subtypes of cognitive decline and spatiotemporal neuropathology progression.^8^ Co-pathologies resulting from vascular disease are also common and contribute to cognitive decline in possibly the majority of AD-diagnosed patients.^9^ Cognitive decline rates, the sequence of biomarker abnormality events, and the pattern of brain neurodegeneration can all differ between AD patients.^8,10,11^ Recent studies have shown that even if AD patients have similar disease severity at baseline, over follow-up these patients may have different rates and trajectories of cognitive decline.^12–15^ AD staging and trajectory prediction models based on cognition and AD biomarkers are currently being developed for enriching trial cohorts with subjects whose forecasted decline trajectories complement trial objectives.^16–18^ However, such tools are not yet widely used because of the extensive validation required and the need for standardized validation procedures.^19^

For the foreseeable future, AD clinical trials cannot avoid treatment-independent heterogeneity in disease progression between subjects. This heterogeneity introduces aleatoric uncertainty into power analysis, as each treatment group will have an unpredictable endpoint variance. T-test power analyses also do not account for the epistemic uncertainty in sample size recommendations introduced by forcing equal variance and normality assumptions. These limitations prevent trial designers from using intrinsically non-normally distributed endpoints. Moreover, it is common practice to transform an outcome metric to make it more normally distributed, for example by taking the difference from baseline.^20^ This practice compresses trial data and discards information.^20^

T-test power analysis also assumes that the measured effect size will be greater than or equal to that anticipated. However, inter-subject heterogeneity injects aleatoric uncertainty in the anticipated mean endpoint difference between treatment groups. This uncertainty could be partially factored in by repeating the t-test power analysis using a range of values for the anticipated endpoint mean difference and pooled variance. However, AD clinical trials do not typically report doing so.

If the t-test assumptions do not hold in a trial’s data, and if the anticipated treatment effect is overestimated when planning the trial, the sample size recommended by a two-sample t-test power analysis could be too small to detect the effect at the desired significance level. A too-small sample size increases Type 2 error risk,^21^ potentially making a trial blind to a genuine treatment effect.

New power analysis methods are urgently needed. These methods should account for inter-subject variability in AD progression, should not rely on normality assumptions, and should factor in aleatoric and epistemic uncertainty. A promising candidate method is power analysis by simulation of clinical trials,^20^ using realistic subject-level AD progression trajectories.

### 1.3. Power analysis by simulation of Alzheimer’s disease progression in clinical trials

Large longitudinal datasets are becoming available on diverse AD cohorts containing participants at different stages of the disease.^22^ With these datasets, one can derive disease progression models that map the trajectories of clinical and biological indicators of AD at the cohort and subject levels.^18^ These AD trajectory maps can then be used to generate subject-level disease trajectories to simulate clinical trials. Power to detect injected treatment effects can then be analyzed across simulations.^23^

AD progression models that capture the disease heterogeneity already exist. For example, recent work by Vogel et al. identified four spatiotemporal patterns of tau spreading in the AD brain, each with different demographic and longitudinal cognitive profiles.^8^ The data-driven SuStain machine learning technique identified three spatiotemporal progression patterns of neurodegeneration from brain magnetic resonance images (MRIs) of AD patients.^24^ A study modeling the clinical progression of AD for the goal of symptom prediction identified up to three trajectory classes (stable, slow decliner, or fast decliner) on different cognitive assessments in the Alzheimer’s Disease Neuroimaging Initiative (ADNI) dataset.^25^

While these and similar trajectory subtyping methods have deepened our understanding of AD, such methods group subjects into distinct trajectory classes only at the cohort-level. For clinical trial simulation, disease progression models that map and can generate both cohort- and subject-level trajectories are needed.

Mixed effect models can solve this problem by regarding each subject as a series of observations following a trajectory that deviates slightly from that of the cohort.^26^ A 2019 study by Iddi et al. used a Bayesian mixed effect model to track cognitive decline at the cohort and subject level in ADNI and predict future decline in new subjects.^27^

Recently, several simulation-based power analysis software and programming frameworks have been developed to evaluate studies where mixed effect models are used to analyze data.^28–30^ However, applications of these tools are focused primarily on fields outside medicine. Furthermore, custom model specification is limited, and sampling parameters cannot easily be specified (e.g., number of observations per subject, observation spacing, dropout rate, etc.). These issues limit the generalizability of these tools, making them difficult to use in trial simulation and power analysis in AD and other progressive diseases.

However, a 2012 study by Rogers et al. proposed a Bayesian mixed effect drug-disease-trial model to track AD progression as well as treatment and placebo response in marketed therapeutics.^23,31^ Model parameters were estimated using literature metadata and data from the Critical Path for Alzheimer’s Disease Database (CPAD) on AD clinical trials.^23,32,33^ This model, called *adsim*, could be used to simulate Phase 2a and 3 trials in AD with realistic dropout, different designs, symptomatic and/or disease-modifying treatment effects, and sampling schemes. Trials are simulated by pooling synthetic subject-level trajectories of scores on a commonly used cognitive test called the Alzheimer’s Disease Assessment Scale–Cognitive Subscale (ADAS-Cog).^31,34,35^ Simulated scores are conditioned on treatment effects, subject-level genetic information, and cognition and age at baseline.^31,35^

In 2013, this model was deemed Fit-for-Purpose under the FDA’s Drug Development Tool Qualification Process.^36,37^ To the best of our knowledge, *adsim* is currently the only simulation-based power analysis Drug Development Tool with this qualification. However, the publicly available model is limited: only data from mild and moderate AD dementia patients were used.^23^ Recent clinical trials of AD treatments include people at other stages of the disease. For example, the recent Phase 3 trials of aducanumab and lecanemab included people with AD-related mild cognitive impairment or mild AD dementia.^38–40^ Power analysis results from the public *adsim* do not generalize to these or other trials targeting earlier AD stages. The model uses only the ADAS-cog score as an endpoint and contains strong statistical assumptions; the model’s posterior distribution is sampled directly to simulate subject-level trajectories.^35^ Although the modeling assumptions may be justifiable, they compress information and potentially ignore latent effects of unmeasured disease processes. By using the model to directly simulate data for new subjects, epistemic uncertainty is injected into the power analysis results. The potential impact of this uncertainty on sample size recommendations remains unaccounted for in *adsim*.

An alternative approach is to simulate clinical trials using real subject data directly. A 2021 study by Chen et al. used real AD patient data from the OneFlorida dataset to simulate different trial designs for the drug donepezil.^41^ The objective was to determine whether it was feasible to evaluate donepezil’s safety profile by simulating the drug’s clinical trial using real-world data on subjects who matched the original trial’s inclusion criteria. The study compared serious adverse event rates in the real trial with those in simulated trials comprised of data on real subjects who had been prescribed the same dosages as in the original trial. Simulations had realistic subject randomization, dropout, and a proportional sampling approach to account for the original trial’s race distribution. Simulations had similar adverse event rates to the original trial, demonstrating the feasibility of using real-world data for simulation-based safety evaluation in clinical trials.

However, this study did not test the simulation approach for evaluating the trial’s power to detect donepezil’s cohort-level symptomatic effect on cognition. The simulations are also retrospective and rely on treatment effects and adverse events already present in the data, thus preventing the prospective generalizability to new trials where effects can only be hypothesized. Also, only one sample of subjects from the OneFlorida dataset was used to simulate one trial. This sampling approach does not factor in the impact of variable cohort composition on trial results. Bootstrap resampling was only done to obtain 95% confidence intervals on the mean simulated adverse event rate. Moreover, the proportional sampling focused only on race and did not replicate other cohort features such as the distribution of AD stage at baseline, despite the inclusion criteria covering moderate to severe AD.^42^

Other recent studies have begun using real-world data to simulate clinical trials longitudinally. A 2021 study by Jutten et al. used real-world data on subjects from ADNI who matched the Phase 3 aducanumab trials’ inclusion criteria to provide an exemplar Phase 3 trial in AD with cognitive test scores as outcome measures.^43^ Commonly used cognitive tests in AD clinical trials are: the Mini Mental State Examination (MMSE),^44^ different subscales within the Alzheimer’s Disease Assessment Scale–Cognitive Subscale (ADAS-Cog),^34^ and the Clinical Dementia Rating–Sum of Boxes (CDRSB).^45^ In the Jutten et al. study, the objective was to calculate the range of treatment effect sizes that could be measured when no effect is actually present. The authors first used a linear mixed effect model to track cohort- and subject-level MMSE, CDRSB, and ADAS-Cog-13 score trends in the selected cohort. For subjects without an 18-month assessment (the Phase 3 aducanumab trials’ duration),^38,39^ those subjects’ own real-world score trajectories were used to calculate test score values at 18 months. The difference from baseline values (e.g., change from baseline in CDRSB) were then computed and used as endpoint metrics in 10,000 simulated trials. To create each simulation, subjects’ scores at 18 months were resampled and randomized 1:1 to groups labelled “treated” and “placebo.” By computing the null treatment effect sizes in each simulation as the between-group difference in means for the change-from-baseline scores, the study obtained the 95% range of observed null effect sizes for each endpoint.

Results showed that group differences in almost all recent real-world anti-amyloid trials fall within the 95% range of null effect sizes in the simulations. This result suggests that Type 1 errors may pose a significant risk for clinical trials in AD. Subgroup analyses with reduced sample sizes could lead to starker imbalances in cognitive decline trajectories between treatment groups, potentially exacerbating the Type 1 error risk.

Developers of new treatments for AD and potentially other progressive diseases might seek to reduce Type 1 error risk by first identifying the range of treatment effect sizes that lie outside the 95% range of potential null effect sizes for the target population. Any effect size outside that range measured in a real-world clinical trial would thus have a low probability of having occurred due to chance imbalance in disease progression between treatment groups. Power analysis could be enhanced if it could also estimate the null treatment effect size range. Information about the null effect size distribution would also likely impact the drug development stages that occur before clinical trials.

To address the limitations of conventional power analysis and those of newer approaches, we propose a novel framework that combines clinical trial simulation using real-world data with disease progression modeling. Our framework can simulate trials of new, disease-modifying treatments of progressive diseases – we show an example from AD below. These simulated trials can then be used to analyze power in different trial designs while accounting for aleatoric and epistemic uncertainty. Our framework can also be used to estimate the null treatment effect size range and Type 1 error rate in a target population.

## 2. Objectives

We introduce our framework as a four-stage procedure. We then answer specific power analysis questions about two recent large-scale clinical trials in Alzheimer’s disease: first, the Phase 3 aducanumab trials EMERGE (NCT02484547) and ENGAGE (NCT02477800);^38,39^ second, the Phase 3 lecanemab trial Clarity AD (NCT03887455).^40^ Our Primary Objectives are: (1) to describe the four stages of our power analysis procedure, focusing on EMERGE and ENGAGE in an example; (2) to analyze power in trials similar to EMERGE and ENGAGE; and (3) to compare our power results with those reported in the published EMERGE and ENGAGE Statistical Analysis Plans.^38,39^ Our Supplementary Objectives are: (1) to analyze power in simulated trials similar to Clarity AD, and (2) to compare our results to those reported in the published Clarity AD results paper.^46^

## 3. Methods

### 3.1. Summary of the power analysis procedure

Our procedure simulates clinical trials by generating subject-level trajectories for a target trial’s endpoint using a mixed effect model of disease progression as well as longitudinal data from real patients who match the trial’s inclusion criteria. We inject a cohort-level treatment effect that alters the rate of disease progression in simulated subjects that are randomly assigned to a treatment group. We use proportional sampling of baseline data from real patients to replicate the baseline disease stage distribution observed in the real-world trial targeted by our power analysis. With this technique, we simulate thousands of trials, each containing a different set of disease progression trajectories but the same cohort-level treatment effect, observation scheme, and proportional sampling. “Power” is the proportion of simulations where a statistically significant treatment effect is detected using a linear mixed effects model. We use this approach to analyze power with 10,000 simulated trials. A flowchart describing the sequence of stages is shown in Figure 1. Each stage is described in detail below.

**Figure 1.**
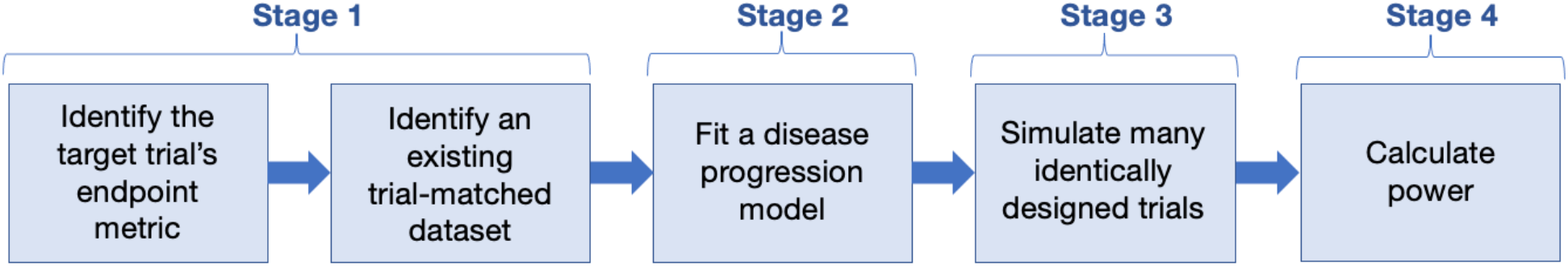
Flowchart describing the sequence of four stages in our power analysis procedure.

### 3.2. Stage 1: Identify the target trial’s endpoint metric and an existing trial-matched dataset

First, we identify the target trial’s primary endpoint. In our Phase 3 aducanumab trial example, the endpoint is the change from baseline in the cognitive test score Clinical Dementia Rating–Sum of Boxes (CDRSBΔbl).^38,39^ We identified this endpoint in the identical analysis protocols outlined in the EMERGE and ENGAGE Statistical Analysis Plans (SAPs) that are publicly available on ClinicalTrials.gov.^38,39^

Next, we identify a longitudinal dataset on real-world subjects who match the target trial’s inclusion criteria and where endpoint observations are available on each subject. Subject data used in the preparation of this article were obtained from the Alzheimer’s Disease Neuroimaging Initiative (ADNI) database (https://adni.loni.usc.edu), specifically from cohorts ADNI1, ADNIGO, ADNI2, and ADNI3.^47–50^. The ADNI was launched in 2003 as a public-private partnership, led by Principal Investigator Michael W. Weiner, MD. The primary goal of ADNI has been to test whether serial magnetic resonance imaging (MRI), positron emission tomography (PET), other biological markers, and clinical and neuropsychological assessment can be combined to measure the progression of mild cognitive impairment (MCI) and early Alzheimer’s disease (AD). For up-to-date information, see www.adni-info.org. The ADNI study received ethics approval from all participating institutions and informed consent from all participants before inclusion in the study.

We selected subjects from ADNI who fit our target trials’ key inclusion criteria at baseline: i.e., subjects meet the clinical criteria for AD-related mild cognitive impairment (MCI) or mild AD dementia, are between 50 and 85 years old, have a CDR score of 0.5 and an MMSE score between 24 and 30, are APOE genotyped, and are positive for abnormal amyloid levels.^38,39^ We determined amyloid status by using ADNI-reported positron emission tomography tracer measures of brain amyloid levels at baseline (PiB SUVR > 1.2 or AV45 SUVR > 1.11),^51,52^ or cerebrospinal fluid measures of amyloid-beta 42 at baseline (Aβ42 ≤ 980 pg/ml).^53^

Out of 2,420 available subjects in the described ADNI datasets at the time of download, 1,040 subjects met our criteria for abnormal amyloid levels. Of these abnormal amyloid positive subjects, 563 (494 MCI, 69 mild AD dementia) met the EMERGE and ENGAGE inclusion criteria, with an average of approximately 5.4 observations per subject and an average follow-up time across subjects of approximately 4 years.

### 3.3. Stage 2: Fit a disease progression model to the trial-matched data

Here we fit a time series model to our trial-matched dataset to track the cohort- and subject-level trajectories of the target trial’s primary endpoint. This model can be relatively simple. Our objective is to model the observed endpoint trajectories in a manner that lets us replicate or interpolate those trajectories across sets of observation times that were not present in the original data. This step is key for simulating clinical trials in Stage 3, where endpoint values are calculated at arbitrary observation times for simulated subjects (discussed in detail in Section 3.4 below).

To map trajectories in our aducanumab example, we use a linear mixed effect model that nearly matches the slope analysis model in the EMERGE and ENGAGE SAPs.^38,39^ We use restricted maximum likelihood to fit the model to our Stage 1 trial-matched dataset using the *lmer* function in the *lme4* package in *R*.^54^ The outcome metric is CDRSBΔbl. The fixed effects are median-centred baseline CDRSB and Mini Mental State Examination (MMSE)^44^ scores, years from baseline, APOE4 status (carrier or not),^55^ symptomatic AD medication use at baseline (yes or no), and an interaction term for median-centred CDRSB with years from baseline. Increasing CDRSBΔbl scores indicate worsening cognitive impairment.^56^ Random effects by subject are defined for years from baseline in order to obtain subject-specific slope parameters. We use all available data on trial-matched subjects to fit the model (proportional sampling is only done in simulated trials in Stage 3). The *lmer* model syntax is thus:

~~~
MCI_mild_AD_Mdl_diff_mc <- lmer(diff_bl_CDRSB ∼ 0 +
 CDRSB_bl_mc +
 CDRSB_bl_mc:Years_bl +
 Years_bl +
 MMSE_bl_mc +
 APOE4_status +
 ADmeds_status +
 (0 + Years_bl | RID),
data = MCI_mild_AD_data_Abpos)
~~~

Figure 2 shows CDRSBΔbl values over time in our trial-matched dataset, where the cohort trend is shown in red, and the observations of one exemplar subject are shown in green.

**Figure 2.**
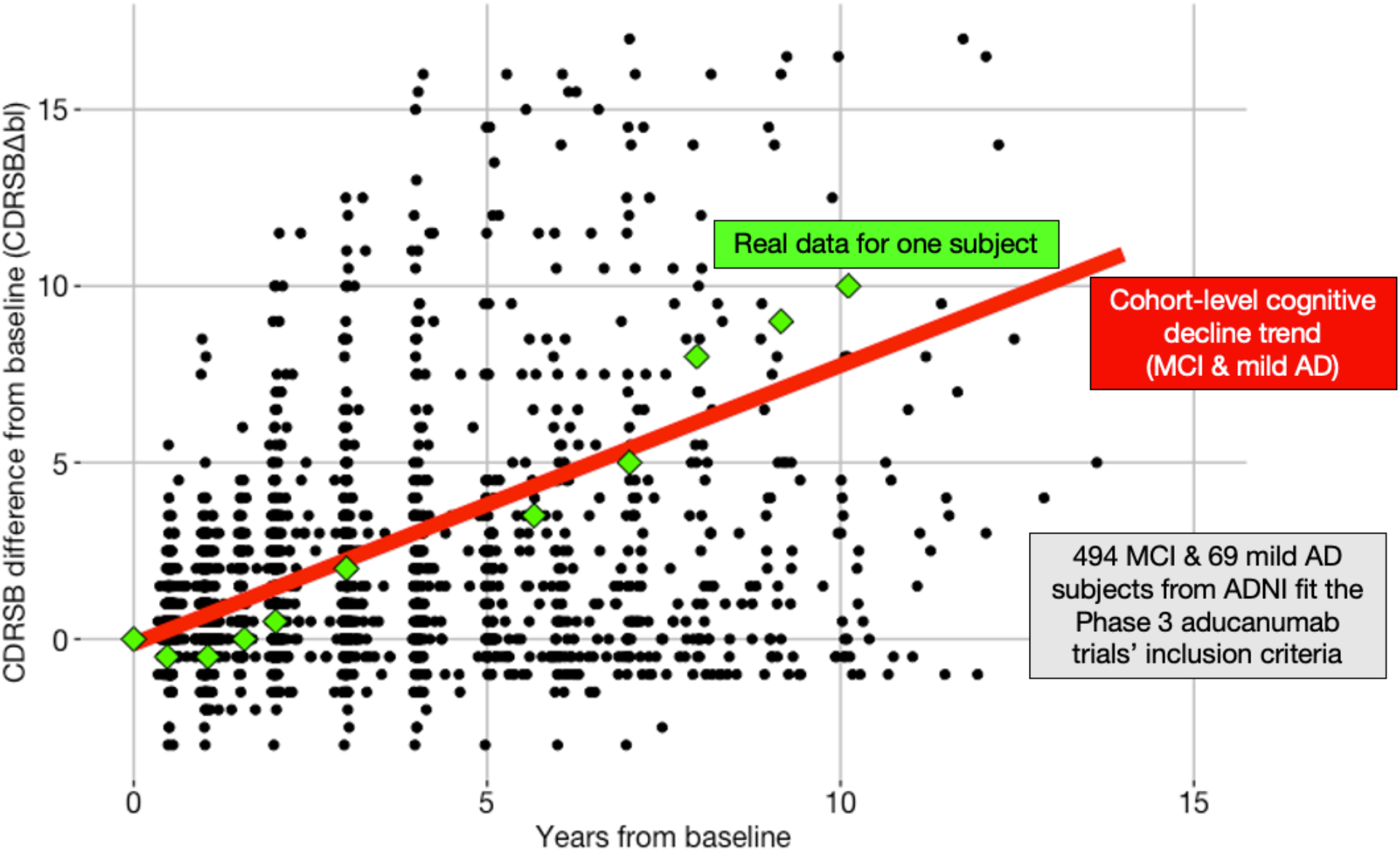
CDRSBΔbl scores over time in our trial-matched dataset from ADNI1, ADNIGO, ADNI2, and ADNI3. 563 subjects fit the inclusion criteria for the Phase 3 aducanumab trials. 494 subjects met clinical criteria for AD-related MCI and 69 met criteria for mild AD dementia. The red line shows a cohort-level CDRSBΔbl trend over years from baseline (generated using the cohort-level predictions of the Stage 2 model, holding other fixed effect variables constant).^57^ Green diamonds show exemplar data for one subject.

After fitting this model, we referred to the Nakagawa *R*^2^ for mixed-models as an absolute value goodness-of-fit measure.^58,59^ With the above model we obtain a conditional *R*^2^ of 0.912, which can be conceptualized as the proportion of the variance in the data being explained by the fixed and random effects in the model.^59^

Importantly, with this trajectory model example we are not advocating any particular procedure for variable or model selection. Our intent with referencing our model’s Nakagawa *R*^2^ is to indicate that the model adequately maps the cohort- and subject-level trajectories. Any appropriate model selection procedure may be used.

The next step is to save the trial-matched dataset along with the model parameters relevant to Stage 3. The saved parameters are: (1) the fixed effects coefficients and their standard errors; (2) the subject-specific random effect estimates that describe subject-specific differences from the cohort-level fixed effect slope coefficient, along with the subject-specific standard errors on those random effects; and (3) the standard deviation of the residuals. These parameters and the trial-matched data will be used to simulate clinical trials in Stage 3 using a data-generating equation that matches the structure of the Stage 2 model.

### 3.4. Stage 3: Simulate many identically designed clinical trials

#### 3.4.1. Define trial sampling parameters, treatment allocation, and significance level

For the EMERGE and ENGAGE trials, the ratio of MCI to mild AD dementia was approximately 4:1.^2^ In our present example we set the number of subjects per treatment group at baseline to 535 subjects (combined total = 1,070), as recommended by the SAP power analysis. EMERGE and ENGAGE had 4 analysis visits spaced approximately 26 weeks apart, and subjects were randomized 1:1 to treated and placebo groups.^2,38,39^ Analysis visits are defined in the SAPs as the observation times in the trial that are to be used in the treatment efficacy analysis. We also encode dropout in our simulations. We define dropout as the percentage of subjects who begin the trial but are not observed at the final analysis visit. As in the EMERGE and ENGAGE power analysis, we define a dropout of 30%.^38,39^ We define a disease-modifying treatment effect as a 25% percent reduction in the cohort-level rate of CDRSBΔbl increase. Our significance level in this example is set by a two-sided α = 0.05. In all examples and experiments below, there is no dose stratification – we compare a single treatment group with a single control group.

#### 3.4.2. Define the data-generating equation

Here we define the equation that will generate cohort- and subject-level cognitive decline trajectory data in our simulated clinical trial. The equation has the same structure as the Stage 2 trajectory model:

~~~
diff_bl_CDRSB_simu =
 (beta_1 + beta_1_delta)*CDRSB_bl_mc_jit +
 (beta_2 + beta_2_delta + gamma_1 + gamma_1_delta)*Year_jit +
 (beta_3 + beta_3_delta)*MMSE_bl_mc_jit +
 (beta_4 + beta_4_delta)*APOE4_status +
 (beta_5 + beta_5_delta)*ADmeds_status +
 (beta_6 + beta_6_delta)*CDRSB_bl_mc_jit*Year_jit +
 (beta_7 + beta_7_delta)*treated_status*Year_jit +
 error
~~~

Parameters and subject-specific baseline data from Stage 2 are input to this equation to simulate (i.e., calculate) endpoint values, `diff_bl_CDRSB_simu`, at arbitrary user-defined years from baseline observations times, `Year_jit`. We calculate values for `Year_jit` using the user-defined number of analysis visits and the visit spacing. Our simulated visits correspond to the Baseline, Week 26, Week 50, and Week 78 analysis visits in EMERGE and ENGAGE.^38,39^

We jitter each subject’s observation time values to simulate subjects being observed within an analysis visit window (hence the `_jit` on `Year_jit`). Jittering matches the window in the real trials of approximately ±13 weeks around each analysis visit.^38,39^ While we could simulate any distribution of observation time jitter (e.g., gaussian or other), in our present example we sample jitter values from a continuous uniform distribution with an interval of approximately ±13 weeks.

The `beta_i` terms in the data-generating equation are the coefficients of the Stage 2 model fixed effect variables. The variables in the data-generating equation being multiplied by the `beta_i` represent the fixed effect variables in the Stage 2 model. The `beta_i_delta` terms correspond to error values for the corresponding `beta_i`. Those error values are sampled from a mean-zero normal distribution with standard deviation equal to the standard error of the corresponding `beta_i`.

The `gamma_1` term is the subject-specific difference value from the cohort-level slope, `beta_2`, which represents the cohort-level rate of cognitive decline in the model. The `gamma_1_delta` term corresponds to a subject-specific error value for a subject’s `gamma_1`, sampled from a mean-zero normal distribution with standard deviation equal to the standard error of the subject’s `gamma_1`.^60^ Together, `beta_2 + beta_2_delta + gamma_1 + gamma_1_delta` describe the treatment-independent subject-specific slope, i.e., the subject-specific rate of cognitive decline.

To inject a slope-altering (i.e., disease-modifying) effect for treated subjects, we add a treatment effect term, `treated_status`. This term indicates whether a given subject has been assigned to the treatment or control group. If the subject is treated, then `treated_status` is set to 1, and an amount `beta_7` is added to the subject’s slope. The `beta_7` term encodes a slope-altering treatment effect as a user-defined percent reduction in the cohort-level rate of CDRSBΔbl increase, i.e., a percent reduction in `beta_2`. In our example, this user-defined percent reduction is set to 25%, as in the EMERGE and ENGAGE SAPs’ power analysis.

The `beta_7_delta` term corresponds to a subject-specific shift in the cohort-level treatment effect term. Shift values are sampled from a mean-zero normal distribution with a user-defined standard deviation (here we arbitrarily set the standard deviation to 0.05 units). To better model between-subject differences in treatment effect, other distributions could be used, including ones conditional on a subject’s clinical and/or biological characteristics at baseline and known decline trajectories.

Finally, the `error` term corresponds to an observation-specific error value for the calculated endpoint. Values for this term are sampled from a mean-zero normal distribution with standard deviation equal to that of the residuals in the Stage 2 model.

#### 3.4.3. Synthesize new subjects and randomize them to treated and control groups

With the above parameters defined, we sample with replacement the trial-matched subjects’ baseline data, where each sample is paired with its corresponding subject-specific trajectory parameters, i.e., the `gamma_1` values with their standard errors. This resampling process is constrained to preserve the distribution of AD stage at baseline in EMERGE and ENGAGE, i.e., 4:1 MCI to mild AD dementia.

For each resampled subject, we jitter the baseline median-centred CDRSB and MMSE scores by adding to each score a randomly sampled value corresponding to measurement error defined as a single step under a clinically significant score change (hence the `_jit` label on `CDRSB_bl_mc_jit` and `MMSE_bl_mc_jit`). For both CDRSB and MMSE, subclinical error in this context has a magnitude of at most 0.5.^61^ We sample a jitter value from a vector containing values –0.5, 0, and 0.5, then add the sampled value to the corresponding `CDRSB_bl_mc` or `MMSE_bl_mc` score. Each value has equal probability of being sampled, and values are sampled separately for CDRSB and MMSE. As a result, approximately 33% of subjects will not have CDRSB or MMSE jittered, and approximately 11% of subjects will have neither score jittered. Each subject will nonetheless have a unique cognitive decline trajectory because of subject-specific `gamma_1_delta` term and, in treated subjects, the `beta_7_delta` term.

Resampling and jittering each real subject’s baseline data in this way allows us to increase the sample size of our simulated trial beyond that available in the trial-matched dataset. This approach preserves the cohort-level trajectory statistics while providing synthetic subjects.

Next, we randomize our subjects 1:1 to treated and control groups by randomly setting the value of each subject’s `treated_status` variable to 0 or 1.

#### 3.4.4. Calculate endpoint values at each observation time using the data-generating equation

Finally, we input each synthetic subject’s baseline data, `Year_jit` vector, and other cohort- and subject-level parameters into the data-generating equation. We then calculate a `diff_bl_CDRSB_simu` score at each observation time for each synthetic subject. We restrict these scores to the range –18 to +18 (the range of real-world CDRSBΔbl scores).^62^ We also round simulated scores to the nearest 0.5 decimal value (e.g., simulated scores 2.3 and 2.7 are rounded to 2.0 and 3.0, respectively).^62^ To encode dropout over time, at each analysis visit all subsequent observations are deleted for approximately 11.2% of subjects. By deleting these future observations, only 70% of subjects complete the trial, signifying 30% dropout by the final analysis visit. The process described here and above simulates a single clinical trial and is illustrated in Figure 3.

**Figure 3.**
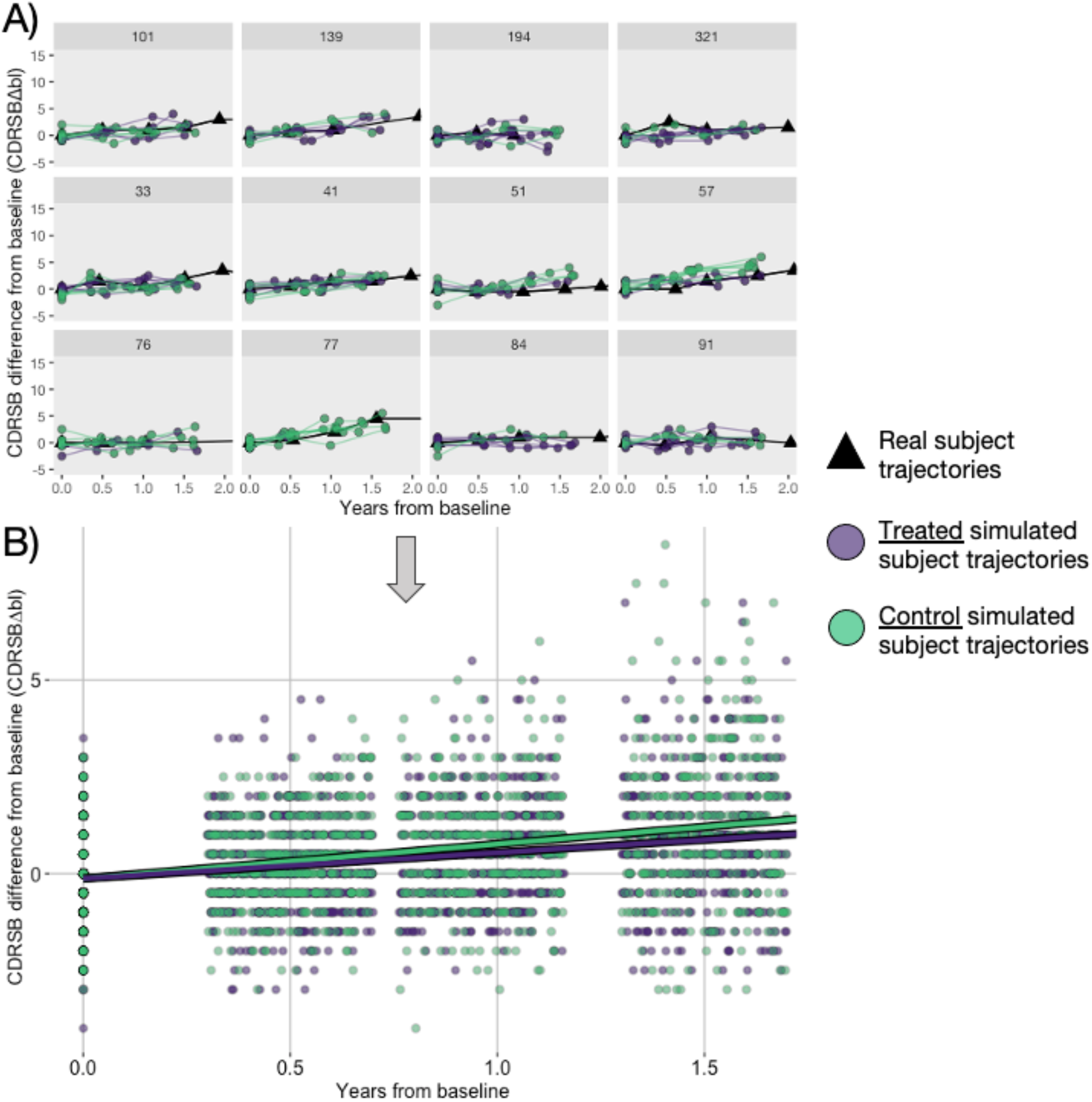
An illustration of how a single clinical trial is simulated. A) Each panel shows a real-world CDRSBΔbl trajectory for a real ADNI subject together with the trajectories of synthetic subjects that are derived from the panel’s corresponding real trajectory. Panel titles indicate the real-world subjects’ anonymized ADNI identification numbers. To generate the synthetic trajectories, baseline data from the corresponding real subject are sampled with replacement then jittered before being input to the data-generating equation. Synthetic CDRSBΔbl scores are generated at each jittered observation time and are rounded to the nearest 0.5 decimal value. Black lines and triangles indicate trajectories of real subjects selected from ADNI. Coloured lines and dots indicate trajectories of simulated subjects. Purple indicates a treated subject and green a control. B) Trajectories of simulated treated and control subjects (here, 535 per group) are pooled into one dataset as a simulated clinical trial. Analysis visits occur at baseline, Week 26, Week 50, and Week 78. Each analysis visit after baseline occurs within a visit window, illustrated in B as approximately ±10 weeks to highlight observation clustering around each analysis visit. Note that visit windows are ±13 weeks in the Section 3.6 EMERGE and ENGAGE simulation Experiments, as in the real-world trials. Respectively, the purple line and the green line represent the group-level trajectories of treated and control groups. These group-level trajectories diverge over the duration of the trial because of the injected disease-modifying treatment effect: a 25% reduction in the rate of CDRSBΔbl increase for treated subjects vs. controls. By the end of the trial, the treated group has a lower average CDRSBΔbl compared to controls, indicating less cognitive impairment. Note that synthetic CDRSBΔbl scores at baseline are distributed around zero because of the observation-level error term, the fixed effect coefficient error terms, and baseline data jittering in the data-generating equation.

#### 3.4.5. Simulate 10,000 clinical trials

We now simulate 10,000 trials by repeating the procedures in Sections 3.4.3 and 3.4.4 10,000 times. Each simulation will contain a different set of cognitive decline trajectories because of the unique sampling of subjects, and the jitter, `_delta` and `error` term values input to the data-generating equation. However, the total number of subjects at baseline and the distribution of subjects’ AD stage at baseline (4:1 MCI to mild AD dementia) remains the same between simulations, as do all `beta_i`. The trial’s number of analysis visits, the visit spacing and window, and the dropout rate also remain the same. Our set of 10,000 simulated trials here is analogous to running 10,000 real-world clinical trials with the same design, where each trial has a different sampling of subject-level decline trajectories drawn from the trial’s target population. We can then calculate statistics across these simulated trials, including power.^43,63^

### 3.5. Stage 4: Calculate power

In each of the 10,000 simulated trials, we fit a linear mixed effect model across the simulated observations to estimate the cohort-level treatment effect size. In this model, the simulation-specific treatment effect size is the coefficient of the model’s `treated_status:Year` term, which signifies the group difference in slope for treated subjects vs. controls. This linear model has the same structure as the data-generating equation and is fit to the simulated trial data using restricted maximum likelihood using *lmer* in package *lme4* in *R*:

~~~
simModel <- lmer(diff_bl_CDRSB_simu ∼ 0 +
 CDRSB_bl_mc +
 Year +
 MMSE_bl_mc +
 APOE4_status +
 Admeds_status +
 CDRSB_bl_mc:Year +
 treated_status:Year +
 (0 + Year | subj_id),
data = simData)
~~~

We define “power” as the proportion of the 10,000 simulations where a statistically significant treatment effect size was detected with a two-sided α under 0.05 using the above model. Statistical significance is indicated if the 95% confidence interval on the `treated_status:Year` coefficient excludes zero. The interval was defined as ±2 standard errors on the coefficient, following recommendations in Gelman and Hill.^64^ In our example trial with the parameters specified in Section 3.4.1, we obtain a statistically significant estimate for our treatment effect in 7,185 out of 10,000 simulations, signifying 71.85% power.

By calculating power for a range of sample sizes (i.e., by repeating Stages 3 and 4 using a range of values for the total number of subjects), we can generate a power curve that lets us determine the number of subjects needed to detect our 25% treatment effect at a given power and significance level. Note that uncertainty intervals can be calculated around the estimated required sample size by obtaining additional samplings, for example by repeating Stage 3 multiple times with the same trial sampling parameters and significance level.

Our output number of subjects recommendation is analogous to the t-test power analysis sample size recommendation. However, our approach is more versatile because we can evaluate power for parameters other than number of subjects, such as number of analysis visits, visit spacing and windowing, dropout rate, and others.

Figure 4A shows power curves for a range of sample sizes and treatment effects in simulated trials similar to EMERGE and ENGAGE. Experiment results are also labelled in Figure 4A and are discussed in Results (Section 4).

**Figure 4.**
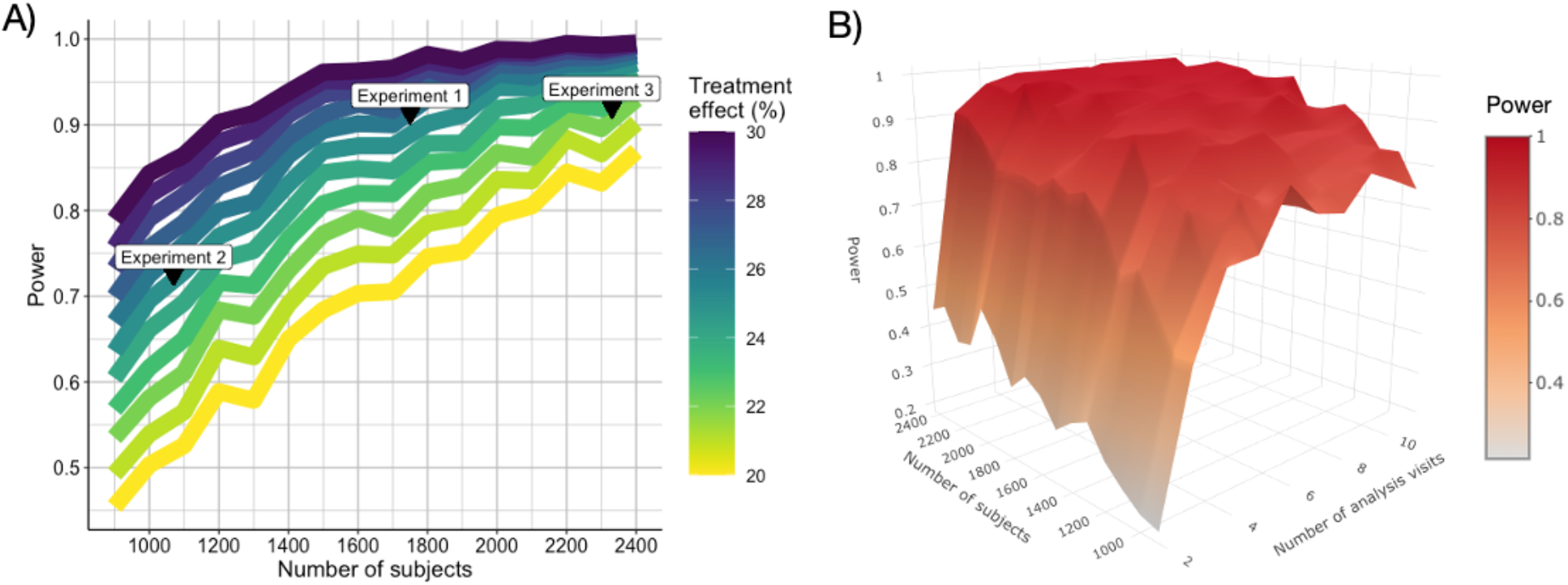
A) Power for simulated trials with the EMERGE and ENGAGE SAP analysis visit scheme but estimated for different treatment effects and numbers of subjects at baseline (combined total for treatment and control groups). Number of subjects ranges from 900 to 2,400 in 100 subject increments. Colours indicate treatment effect, from 20% to 30% in 1% increments, as the cohort-level percent reduction in the rate of CDRSBΔbl increase for treated subjects vs. controls. Power increases with sample size and treatment effect. Labels indicate results for the Experiments (described in Section 4.1). Note that the curves are not perfectly smooth because each point indicates power calculated within a finite set of 1,500 simulations. B) A 3D visualization of a power surface for simulated trials with the same 25% treatment effect, but different combinations of numbers of subjects and numbers of equally spaced analysis visits. The surface is not perfectly smooth because each point indicates power calculated within a set of 150 simulations. Power tends to increase with sample size and number of visits. Number of subjects ranges from 900 to 2,400 in 100 subject increments. Number of analysis visits ranges from 2 to 11 in 1 visit increments. Different combinations of sample size and number of visits may yield similar power. Higher dimensional power surfaces are possible with combinations of more than two trial parameters.

Figure 4B shows a visualization of a 3D “power surface,” which in this case displays power results for different combinations of sample sizes and number of analysis visits. This power surface suggests that 90% power may be achievable in a trial design with 3 visits and approximately 2,300 subjects, or in a trial with 4 visits with approximately 1,800 subjects. Ignoring subject dropout, the first design requires 300 fewer subject-level observations total compared to the second design. Reducing the number of observations could reduce logistical and data processing burdens and could shorten a trial’s duration, thus reducing costs and accelerating treatment development.

The power surface in Figure 4B is essentially the same as the “iso-power contour” proposed by Baker et al.^65^ To introduce the iso-power contour, Baker et al. calculated power in several experimental paradigms and data acquisition modalities (e.g., reaction times, functional MRI, electroencephalography) in psychology and human neuroscience. In each experiment, the authors used different combinations of numbers of subjects and observations per subject. Their study concluded that the number of observations had a significant effect on power in all the studied experimental paradigms.

Expanding on the concept of iso-power contours, we propose the *n*-dimensional power surface, which could indicate power for different combinations of any *n* – 1 trial parameters, for example: different sample sizes, numbers of analysis visits, visit spacings, anticipated dropout rates, cohort- and subject-level treatment effects, analysis visit windows, measurement error, analysis model specifications, and more. Trial designers could then use numerical optimization to identify combinations of trial parameters that complement a trial’s objectives, power targets, and time/economic constraints. We now outline the experiments that target our present study’s Primary Objectives 2 and 3.

### 3.6. Experiments

Each Experiment is presented as a power analysis question. To complete Primary Objectives 2 and 3, we analyze power in 10,000 simulated trials per Experiment using the Phase 3 aducanumab trial-matched dataset, trajectory model, trial simulation, and power analysis procedure described above. The Experiment questions and the simulated results are presented in Section 4.1 below.

To complete our Supplementary Objectives, we used our four-stage procedure to answer a power analysis question about the Phase 3 lecanemab trial Clarity AD in one Experiment with 10,000 simulations of the trial. Lecanemab is a human monoclonal antibody designed to attack soluble amyloid-beta protofibrils in the early-stage AD brain, potentially slowing future cognitive decline.^46^ Clarity AD detected a statistically significant 27% slowing of cognitive decline in subjects treated with lecanemab compared to placebo.^46^ See Supplementary analyses (Section 7) for a detailed description of our power analysis procedure applied here. The Experiment question and results are presented in Section 4.2 below.

## 4. Results

### 4.1. Power analysis results for the Phase 3 aducanumab trials EMERGE and ENGAGE

In Experiment 1 we ask: *How many subjects per group are required to have 90% power at a two-sided α = 0.05 to detect a 25% reduction in the rate of CDRSBΔbl increase?* Here our 10,000 simulations indicate that approximately 875 subjects per treatment group are needed (combined total = 1,750 subjects). This number is 340 subjects per group more than the 535 subjects per group recommended by the t-test power analysis at this power and two-sided α level in the EMERGE and ENGAGE SAPs. This result is labelled in Figure 4A as “Experiment 1.”

In Experiment 2 we ask: *What is the power at a two-sided α = 0.05 to detect a 25% treatment effect if the trial has 535 subjects per group (combined total = 1,070 subjects), as recommended by the EMERGE and ENGAGE SAPs’ power analysis?* We obtained a statistically significant estimate for this treatment effect in 7,185 out of 10,000 simulations, signifying 71.85% power. This power value is 18.15% lower than the 90% power at a two-sided α = 0.05 implied in the SAPs. This result is labelled in Figure 4A as “Experiment 2.”

In Experiment 3 we ask: *How many subjects per group are required to have 90% power at a two-sided α = 0.05 to detect a 22% treatment effect, the same effect that was measured as the statistically significant treatment effect in the EMERGE high dose group?*^2^ Our 10,000 simulations show that approximately 1,165 subjects per group are required (combined total = 2,330 subjects). This number is 630 more subjects per group than the 535 that were recommended by the SAP power analysis. Also, our simulation-recommended number of subjects per group here is at least 617 more than were included in the real-world EMERGE high dose and placebo groups at baseline (547 and 548 subjects, respectively).^2^ This result is labelled in Figure 4A as “Experiment 3.”

### 4.2. Power analysis results for the Phase 3 lecanemab trial Clarity AD

In Experiment S1 we ask: *How many subjects per group are required to have 90% power at a two-sided α = 0.05 to detect a 25% reduction in the rate of CDRSBΔbl increase?* This 25% reduction is the anticipated treatment effect used in the real Clarity AD sample size calculation.^46^ Our 10,000 simulations show that approximately 890 subjects per treatment group are required (combined total = 1,780 subjects). 890 subjects is 107 more than the 783 subjects per group (combined total = 1,566 subjects) recommended by the trial’s original t-test power analysis at this power and two-sided significance level.^46^ This result is labelled in Figure S1 (see Section 7 for Supplementary analyses) as “Experiment S1” on a 25% treatment effect power curve. However, in the real-world trial the target sample size was increased by 200 subjects to account for subjects missing doses due to the Covid-19 pandemic.^46^ This increase brought the new total sample size target to 1,766 subjects. The final total number of subjects who underwent randomization in the trial was 1,795 (898 and 897 subjects to the lecanemab and placebo groups, respectively), which is 15 subjects more than our estimated total number of subjects required to reach 90% power.

## 5. Discussion

### 5.1. Discussion of power results for EMERGE and ENGAGE

In our power analysis of trials similar to EMERGE and ENGAGE, results show that power increases with number of subjects and treatment effect. Our inclusion of realistic inter-subject variability in AD-related progression trajectories of cognitive decline attempts to account for aleatoric uncertainty. This variability increases sample size recommendations beyond those in the published SAPs.

The SAPs’ power analysis may also have used an overestimated anticipated endpoint mean difference value without adequately considering uncertainty. The statistically significant difference in mean CDRSBΔbl between treatment groups measured in EMERGE was 0.39.^2^ This value is 0.11 units less than the SAPs’ anticipated difference in means value of 0.5 CDRSBΔbl points, which we also used in our Section 1.1 t-test power analysis example. The real-world trials also increased their target sample size from 450 to 535 subjects per group based on an interim analysis of available trial data. However, our Experiments’ 1 and 2 results suggests that 535 subjects per group was still insufficient to reach 90% power. The t-test power analysis assumption that the measured effect size will be greater than or equal to that anticipated did not hold in the Phase 3 aducanumab trial data. The SAPs’ power analysis did not account for endpoint mean difference uncertainty and could not consider the impact of inter-subject variability on sample size recommendations. It is therefore possible that the SAP power analyses recommended sample sizes that were too small to detect both the anticipated and measured treatment effect sizes at 90% power at a two-sided α = 0.05. Insufficient sampling during the trial recruitment stage could partly explain the detection of a statistically significant effect for aducanumab being detected only in EMERGE.

### 5.2. Discussion of power results for Clarity AD in the context of EMERGE and ENGAGE

Our Experiment S1 result suggests that the Phase 3 lecanemab trial Clarity AD was adequately powered. Furthermore, our result suggests that this trial required a total number of subjects that was approximately 30 subjects more than our estimated number for the Phase 3 aducanumab trials to detect the same cohort-level 25% treatment effect at 90% power. This relatively small difference in our estimated required sample sizes for these trials could partly be explained by differences between the design of Clarity AD and the identically designed EMERGE and ENGAGE. For example, the Clarity AD inclusion criteria identified subjects with memory impairment specifically in addition to global cognitive impairment,^40^ while EMERGE and ENGAGE focused on global cognitive impairment.^38,39^ The proportions of AD-related MCI and mild AD dementia subjects and the age range of subjects also differed between the described lecanemab and aducanumab trials. Also, EMERGE and ENGAGE had 4 analysis visits and anticipated an overall 30% dropout. Clarity AD had 7 visits and anticipated a dropout of 20%. These cohort and design differences are factored into our simulated trials and our power analysis results. The combined effect on power of different trial design and cohort characteristics for Clarity AD compared to EMERGE and ENGAGE could partly explain our larger estimated number of subjects required for Clarity AD to detect our simulated 25% treatment effect.

### 5.3. Contributions, limitations, and future work

We have proposed a novel power analysis framework that uses disease progression modeling and data from real-world subjects to simulate clinical trials in AD using a cognitive test score as an endpoint. We showcased the procedure by analyzing power in two recent large-scale clinical trials in AD. By using efficient coding practices and parallel computing, each set of 10,000 simulations finished in under 18 minutes on a 2020 quad-core Intel i7 13” MacBook Pro.

We describe our method as a four-stage procedure to facilitate the generalizability to different trajectory modeling approaches, different clinical or biological endpoints in different trials or trial phases, or potentially to datasets on other progressive diseases such as amyotrophic lateral sclerosis, multiple sclerosis, cancers, and more. We use an easily implementable data augmentation technique to synthesize data and increase the number of subjects available for simulation. By allowing different model specifications and assumptions in principle, our framework has the potential to account for epistemic uncertainty. For example, in trials like Clarity AD we can analyze power to detect the time to clinically significant cognitive decline on the global Clinical Dementia Rating scale.^46^ Future implementations can use a joint modeling approach, for example to analyze power for endpoints on the progression of tau PET ligand uptake and cognitive decline simultaneously in future trials of potentially tau tangle-clearing treatments.

Our framework can also evaluate a trial’s potential Type 1 error rate by setting the treatment effect to zero, similar to the method proposed by Jutten et al.^43^ To the best of our knowledge, no power analysis approach is adapted to generate Type 1 and Type 2 error estimates while also factoring in aleatoric and epistemic uncertainty, and the impact of different combinations of tunable trial parameters. Our power surface and supplementary analysis results suggests that the number of analysis visits may affect trial power. Future work should also evaluate the sensitivity of power to different trial inclusion criteria and the use of different trial-matched datasets within the same disease (e.g., from the CPAD clinical trials in AD database).^33^ Compared to t-tests power analysis methods currently in use, our procedure could help to more accurately evaluate the impact on power of clinical trial enrichment tools such as those predicting disease progression.^17^ Results of our analyses suggest that trials may need to increase sample sizes to be adequately powered, which would increase trial costs. In response, the demand for trial enrichment tools might increase because these tools could help reduce sample sizes while preserving or enhancing trial power.^17^

We generalize the proportional sampling approach proposed by Chen et al.^41^ to AD staging, preserving proportions at the level of individual simulated trials. Given enough real-world data, proportional sampling could feasibly be extended to biometrics and finer demographic categories such as sex and race. The time is right for our technique, given the wealth of data currently available from diverse, large-scale population neuroscience and aging studies such as the UK Biobank^66^ and many others.

However, the current implementation of our framework is not without limitations. First, by not considering a placebo effect, our examples essentially compare the effects of aducanumab and lecanemab against the effect of the different types of care received by individuals in ADNI. Also, the demographic profile in ADNI is not as heterogeneous as that in the Phase 3 aducanumab or lecanemab trials. ADNI contains data on mostly white, well-educated men, and data collection took place only in the United States and Canada. Data for Clarity AD, EMERGE, and ENGAGE were collected at many different sites with diverse subject pools across North America, Europe, and Asia. Our simulations did not factor in regional differences between subjects and thus may have underestimated true variability. Future implementations in AD should use datasets such as CPAD, where data on placebo-controlled trials in diverse cohorts can be used to factor in the impact of placebo and possibly geographical region on subject-level trajectories.^32,33^

Another key limitation is that a trial’s endpoint and inclusion criteria metrics may not be available in existing longitudinal datasets. While future data collection efforts may consider the role datasets could play in clinical trial design, existing data harmonization-imputation methods could be used to impute absent metrics.^67^

Our linear modeling approach may not fully capture the shape of cohort- and subject-level AD progression, which likely impacts power results. However, our approach is describing a best-case scenario for a trial: the model equation that contains the treatment effect matches the equation used to generate the data. Low power in this scenario likely warrants increased sample sizes. Future work will use non-linear modeling approaches and compare the impact on power estimates between model specifications.

Other modeling frameworks such as group-based trajectory modeling may be suitable for capturing distinct trajectory subgroups around a cohort average trend while simultaneously enabling estimation of subject-level trajectories around the subgroup averages.^68^ Future work should incorporate categorical time mixed models of repeated measures (MMRMs) in the power analysis step to better match the types of analyses done in real-world clinical trials.^69^ MMRMs make no assumptions about the endpoint trajectory shape and were used to analyze data in the Phase 3 aducanumab and lecanemab trials. However, by using a continuous time model to estimate the impact of aducanumab on rate of decline in treated subjects, we more closely replicate the aducanumab slope analysis model. To the best of our knowledge, the Phase 3 trials of aducanumab and lecanemab did not report results from any slope analyses. Moreover, in studying disease-modifying treatments for AD or other progressive diseases, we are arguably most interested in a treatment’s impact on the rate of disease progression (measured by a slope model), not a binary effect separating treatments groups at a trial’s final observation.

## 6. Conclusion

We have described the four stages of our power analysis procedure, presented example applications in trials like the recent Phase 3 aducanumab and lecanemab trials, and compared the results of our analyses to the analysis plans and results of these trials. As our approach explicitly accounts for the cognitive decline variability that implicitly affects treatment effect size and trial power, our simulations show that the number of subjects in the Phase 3 aducanumab trials’ SAPs *and* the number included in the real trials may have been insufficient to give 90% power to detect both the anticipated and measured treatment effect sizes. The sample size referenced here is the one recommended by the EMERGE and ENGAGE power analysis, *not* the sample size that remained after the EMERGE and ENGAGE futility declaration.^3^ The futility declaration may have further reduced a sample size that was already too small when the trials began.

In the Phase 3 lecanemab trial Clarity AD, the original sample size recommended by the t-test power analysis may have been insufficient to ensure 90% power. However, the increased sample size used in the real-world trial may have been sufficient. Our analysis of Clarity AD also suggests that trial power may be affected by the number of analysis visits and the characteristics of subgroups within a cohort.

Our procedure addresses the limitations of commonly used t-test power analysis techniques. This contribution has implications for improving sample size calculations in clinical trials in AD and potentially in other progressive diseases. By presenting our approach as a four-stage procedure, we hope to facilitate the generalizability to future clinical trials in AD and other disorders.

## Data Availability

Data used in preparation of this article were obtained from the Alzheimer’s Disease Neuroimaging Initiative (ADNI) database (https://adni.loni.usc.edu). As such, the investigators within the ADNI contributed to the design and implementation of ADNI and/or provided data but did not participate in analysis or writing of this report. A complete listing of ADNI investigators can be found at: http://adni.loni.usc.edu/wp-content/uploads/how_to_apply/ADNI_Acknowledgement_List.pdf To obtain access to ADNI data, we first registered in the online ADNI system and submitted a written data access application to ADNI describing our research project proposal. Our proposal was evaluated by research personnel at ADNI and was approved. Subsequently, we were granted access to ADNI data, which we used in preparation of this article. ADNI data can be accessed by registering and applying for access at https://adni.loni.usc.edu/data-samples/access-data/. To maintain data security and save computer storage space, simulated datasets used in preparation of this article were automatically deleted after each simulation experiment described. However, our simulation experiments can be replicated upon request. Researchers can submit a research proposal to ADNI to apply for access to ADNI data; after receiving access approval from ADNI, the ADNI dataset can be used to generate simulated datasets similar to ours by following the procedure outlined in our submitted article.

## 7. Supplementary analyses

### 7.1. Our four-stage power analysis procedure applied to the Phase 3 lecanemab trial Clarity AD

Here we used our four-stage procedure to simulate the Phase 3 lecanemab trial Clarity AD (NCT03887455)^40^ in an Experiment that answers one power analysis question (defined in Section 4.2). The steps we took within each stage are described below.

#### Stage 1

First we identified the relevant design and power analysis parameters specified in the published Clarity AD results paper.^46^ CDRSBΔbl was the trial’s endpoint, the anticipated treatment effect was a 25% slowing of CDRSBΔbl increase for treated subjects vs. placebo, and the desired power was 90% at a two-sided α = 0.05. The trial had a ratio of approximately 3:2 for MCI to mild AD dementia subjects randomized 1:1 to lecanemab and placebo groups. There were 7 analysis visits spaced approximately 13 weeks apart, and 20% overall dropout anticipated.

#### Stage 2

Here we obtained a continuous-time linear mixed effect model tracking CDRSBΔbl trajectories in real-world data from ADNI subjects who fit Clarity AD inclusion criteria at baseline. Inclusion criteria are listed on ClinicalTrials.gov.^40^ The model here is a continuous-time analogue of the mixed model for repeated measures with categorical-time used in the real trial.^46^ The outcome variable is CDRSBΔbl. The fixed effects are median-centred baseline CDRSB scores, years from baseline, a clinical subgroup indicator (= 0 for MCI or = 1 for mild AD dementia), APOE4 status (carrier or not), symptomatic AD medication use at baseline (yes or no), and an interaction term for median-centred baseline CDRSB with years from baseline. Random effects by subject are defined for years from baseline.

#### Stage 3

In this stage we generated a set of 1,500 simulated trials having the Clarity AD analysis visit scheme. Each simulation had the same 25% treatment effect and the same sample size at baseline = 900 subjects total (treated and control groups combined).

#### Stage 4

Within this set of simulations, we calculated the power to detect the 25% treatment effect.

#### Repeat Stages 3 and 4 to generate Figure S1

We repeated Stages 3 and 4 for 13 more sets of 1,500 simulated trials, but with the baseline sample size changing between sets of simulations. Sample sizes went from 1,050 to 2,850 subjects in increments of 150 subjects. From the resulting curve plotted in Figure S1, we determined that 90% power would be reached with approximately 1,780 subjects (result labelled “Experiment S1” in Figure S1).

#### Repeat Stages 3 and 4 to verify the sample size result

To evaluate and verify the estimated sample size value from Figure S1, we repeated Stages 3 and 4 using 10,000 simulations per set but across a narrower range of baseline sample sizes (e.g., changing from 1,760 to 1,800 subjects total in increments of 10 subjects). This process saved computation time and confirmed that approximately 1,780 subjects total are needed for the trial to reach 90% power. With 1,780 subjects total, we obtained a statistically significant estimate for our 25% treatment effect in 9,048 out of 10,000 simulations, indicating 90.48% power. Future implementations will use more powerful computer hardware to quickly generate power curves like that in Figure S1 but using 10,000 simulations per simulation set with finer increments of number of subjects between simulation sets.

**Figure S1.**
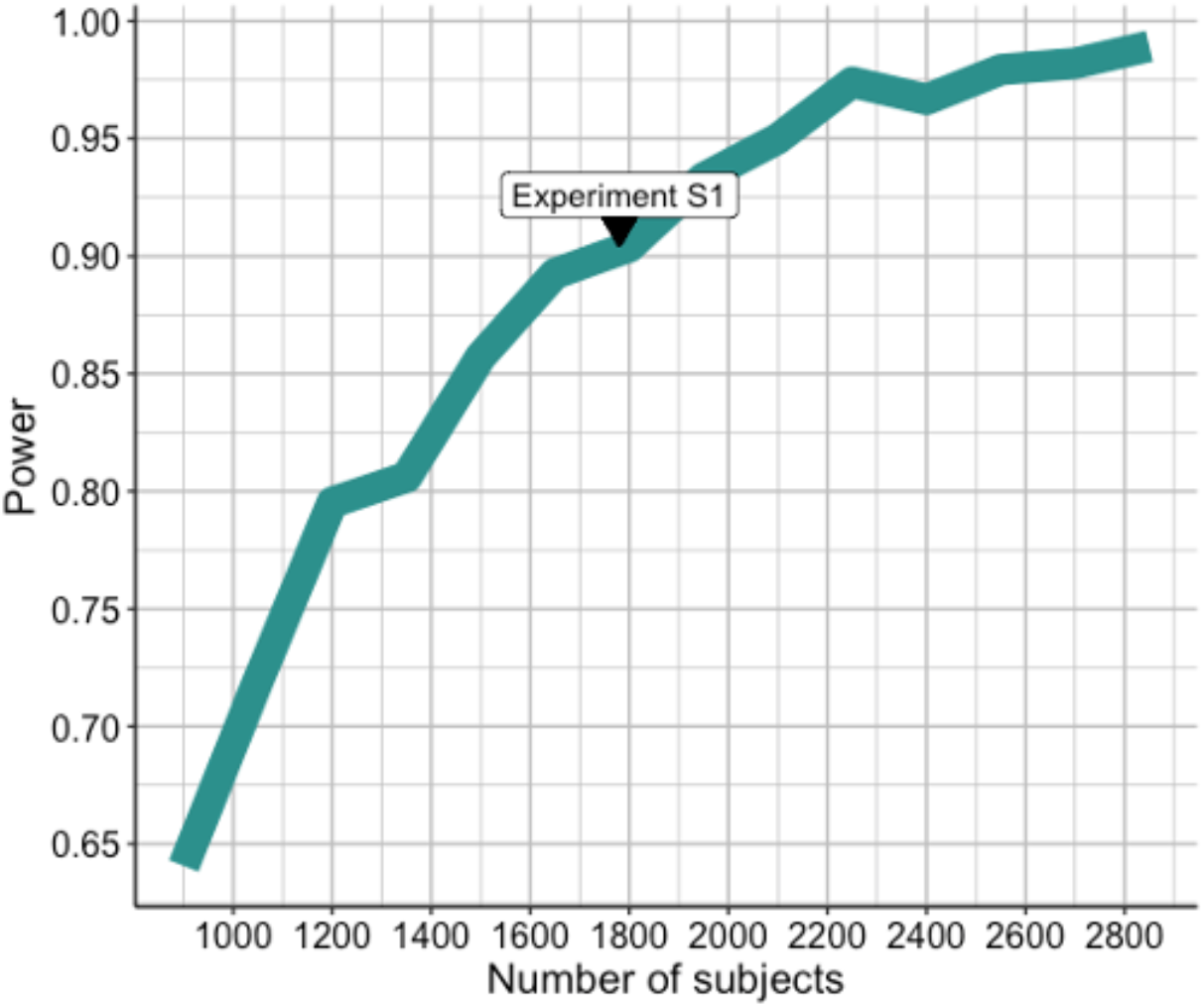
Power for simulated trials with the Clarity AD analysis visit scheme, estimated across a range of baseline sample sizes (total number of subjects for treated and control groups combined). Number of subjects goes from 900 to 2,850 in 150 subject increments. Each simulated trial had the same 25% treatment effect, defined as the cohort-level percent reduction in the rate of CDRSBΔbl increase for treated subjects vs. controls. Note that the curve is not perfectly smooth because each point indicates power calculated within a finite set of 1,500 simulations. Power increases with sample size. The “Experiment S1” label indicates the result for that experiment (described in Section 4.2), where approximately 1,780 subjects total (890 per treatment group) are required for the trial to reach 90% power with a two-sided α = 0.05.

## 8. Acknowledgements

Data collection and sharing for this project was funded by the Alzheimer’s Disease Neuroimaging Initiative (ADNI) (National Institutes of Health Grant U01 AG024904) and DOD ADNI (Department of Defense award number W81XWH-12-2-0012). ADNI is funded by the National Institute on Aging, the National Institute of Biomedical Imaging and Bioengineering, and through generous contributions from the following: AbbVie, Alzheimer’s Association; Alzheimer’s Drug Discovery Foundation; Araclon Biotech; BioClinica, Inc.; Biogen; Bristol-Myers Squibb Company; CereSpir, Inc.; Cogstate; Eisai Inc.; Elan Pharmaceuticals, Inc.; Eli Lilly and Company; EuroImmun; F. Hoffmann-La Roche Ltd and its affiliated company Genentech, Inc.; Fujirebio; GE Healthcare; IXICO Ltd.; Janssen Alzheimer Immunotherapy Research & Development, LLC.; Johnson & Johnson Pharmaceutical Research & Development LLC.; Lumosity; Lundbeck; Merck & Co., Inc.; Meso Scale Diagnostics, LLC.; NeuroRx Research; Neurotrack Technologies; Novartis Pharmaceuticals Corporation; Pfizer Inc.; Piramal Imaging; Servier; Takeda Pharmaceutical Company; and Transition Therapeutics. The Canadian Institutes of Health Research is providing funds to support ADNI clinical sites in Canada. Private sector contributions are facilitated by the Foundation for the National Institutes of Health (www.fnih.org). The grantee organization is the Northern California Institute for Research and Education, and the study is coordinated by the Alzheimer’s Therapeutic Research Institute at the University of Southern California. ADNI data are disseminated by the Laboratory for Neuro Imaging at the University of Southern California.

## References

1. Ard, M. C. & Edland, S. D. Power Calculations for Clinical Trials in Alzheimer’s Disease. J. Alzheimers Dis. 26, 369–377 (2011).

2. Dhillon, S. Aducanumab: First Approval. Drugs 81, 1437–1443 (2021).

3. Budd Haeberlein, S. et al. Two Randomized Phase 3 Studies of Aducanumab in Early Alzheimer’s Disease. J. Prev. Alzheimers Dis. (2022) doi:10.14283/jpad.2022.30.

4. Lakens, D. Calculating and reporting effect sizes to facilitate cumulative science: a practical primer for t-tests and ANOVAs. Front. Psychol. 4, (2013).

5. Jones, S. R. An introduction to power and sample size estimation. Emerg. Med. J. 20, 453–458 (2003).

6. R Core Team. R: A language and environment for statistical computing. https://www.R-project.org/ (2021).

7. Champely, S. pwr: Basic functions for power analysis. https://CRAN.R-project.org/package=pwr (2020).

8. Vogel, J. W. et al. Four distinct trajectories of tau deposition identified in Alzheimer’s disease. Nat. Med. 27, 871–881 (2021).

9. Petrovitch, H. et al. AD lesions and infarcts in demented and non-demented Japanese-American men. Ann. Neurol. 57, 98–103 (2005).

10. 2020 Alzheimer’s disease facts and figures. Alzheimers Dement. 16, 391–460 (2020).

11. Venkatraghavan, V., Bron, E. E., Niessen, W. J. & Klein, S. Disease progression timeline estimation for Alzheimer’s disease using discriminative event based modeling. NeuroImage 186, 518–532 (2019).

12. van Rossum, I. A. et al. Injury markers predict time to dementia in subjects with MCI and amyloid pathology. Neurology 79, 1809–1816 (2012).

13. Vos, S. J. B. et al. Prevalence and prognosis of Alzheimer’s disease at the mild cognitive impairment stage. Brain 138, 1327–1338 (2015).

14. Jack, C. R. et al. Brain beta-amyloid measures and magnetic resonance imaging atrophy both predict time-to-progression from mild cognitive impairment to Alzheimer’s disease. Brain 133, 3336–3348 (2010).

15. Scheltens, N. M. E. et al. Prominent Non-Memory Deficits in Alzheimer’s Disease Are Associated with Faster Disease Progression. J. Alzheimers Dis. 65, 1029–1039 (2018).

16. Zandifar, A. et al. SNIPE score can capture prodromal Alzheimer’s in cognitively normal subjects. bioRXiv (2019) doi:10.1101/541854.

17. Shafiee, N., Dadar, M., Ducharme, S. & Collins, D. L. Automatic Prediction of Cognitive and Functional Decline Can Significantly Decrease the Number of Subjects Required for Clinical Trials in Early Alzheimer’s Disease. J. Alzheimer’s Dis. 84, (2021).

18. Raket, L. L. Statistical Disease Progression Modeling in Alzheimer Disease. Front. Big Data 3, 24 (2020).

19. Cowley, L. E., Farewell, D. M., Maguire, S. & Kemp, A. M. Methodological standards for the development and evaluation of clinical prediction rules: a review of the literature. Diagn. Progn. Res. 3, 16 (2019).

20. Huang, C., Li, P. & Martin, C. R. Simplification or simulation: Power calculation in clinical trials. Contemp. Clin. Trials 113, 106663 (2022).

21. Columb, M. & Atkinson, M. Statistical analysis: sample size and power estimations. BJA Educ. 16, 159–161 (2016).

22. Salimi, Y. et al. ADataViewer: exploring semantically harmonized Alzheimer’s disease cohort datasets. Alzheimers Res. Ther. 14, 69 (2022).

23. Rogers, J. A. et al. Combining patient-level and summary-level data for Alzheimer’s disease modeling and simulation: a beta regression meta-analysis. J. Pharmacokinet. Pharmacodyn. 39, 479–498 (2012).

24. Young, A. L. et al. Uncovering the heterogeneity and temporal complexity of neurodegenerative diseases with Subtype and Stage Inference. Nat. Commun. 9, 4273 (2018).

25. Bhagwat, N., Viviano, J. D., Voineskos, A. N., Chakravarty, M. M., & Alzheimer’s Disease Neuroimaging Initiative. Modeling and prediction of clinical symptom trajectories in Alzheimer’s disease using longitudinal data. PLOS Comput. Biol. 14, e1006376 (2018).

26. Singer, J. D. & Willett, J. B. Applied Longitudinal Data Analysis: Modeling Change and Event Occurrence. (Oxford University Press, 2003). doi:10.1093/acprof:oso/9780195152968.001.0001.

27. Iddi, S. et al. Predicting the course of Alzheimer’s progression. Brain Inform. 6, 6 (2019).

28. DeBruine, L. M. & Barr, D. J. Understanding Mixed-Effects Models Through Data Simulation. Adv. Methods Pract. Psychol. Sci. 4, 15 (2021).

29. Green, P. & MacLeod, C. J. SIMR: an R package for power analysis of generalized linear mixed models by simulation. Methods Ecol. Evol. 7, 493–498 (2016).

30. Kumle, L., Vo, M. L.-H. & Draschkow, D. Estimating power in (generalized) linear mixed models: an open introduction and tutorial in R. Behav. Res. Methods 53, 2528–2543 (2020).

31. Polhamus, D., Rogers, J., Gillespie, B., French, J. & Gastonguay, M. From Evidence Synthesis to Trial Optimization: The adsim Package for Model-based Simulation in Alzheimer’s Disease. in PAGE. Abstracts of the Annual Meeting of the Population Approach Group in Europe. ISSN 1871-6032. (2012).

32. Neville, J. et al. Development of a unified clinical trial database for Alzheimer’s disease. Alzheimers Dement. 11, 1212–1221 (2015).

33. Sivakumaran, S. et al. The Critical Path for Alzheimer’s Disease (CPAD) Consortium: A platform for pre-competitive data sharing, standardization, and analysis to support quantitative tools for AD drug development. Alzheimers Dement. 17, (2021).

34. Kueper, J. K., Speechley, M. & Montero-Odasso, M. The Alzheimer’s Disease Assessment Scale–Cognitive Subscale (ADAS-Cog): Modifications and Responsiveness in Pre-Dementia Populations. A Narrative Review. J. Alzheimers Dis. 63, 423–444 (2018).

35. Polhamus, D. G. & Rogers, J. A. Simulating Clinical Trials in Alzheimer’s Disease. https://bitbucket.org/metrumrg/alzheimers-disease-progression-model-adascog/downloads/adsim_training_doc.pdf (2011).

36. U.S. Food and Drug Administration (FDA). Drug Development Tools: Fit-for-Purpose Initiative. U.S. Food & Drug Administration https://www.fda.gov/drugs/development-approval-process-drugs/drug-development-tools-fit-purpose-initiative (2022).

37. Sinha, V. & Zineh, I. Determination Letter: adsim. U.S. Food & Drug Administration https://www.fda.gov/media/98856/download (2013).

38. Biogen. 221AD301 Phase 3 Study of Aducanumab (BIIB037) in Early Alzheimer’s Disease (ENGAGE): NCT02477800. ClinicalTrials.gov https://clinicaltrials.gov/ct2/show/NCT02477800 (2021).

39. Biogen. 221AD302 Phase 3 Study of Aducanumab (BIIB037) in Early Alzheimer’s Disease (EMERGE): NCT02484547. ClinicalTrials.gov https://clinicaltrials.gov/ct2/show/NCT02484547 (2021).

40. Eisai Inc. & Biogen. A Study to Confirm Safety and Efficacy of Lecanemab in Participants With Early Alzheimer’s Disease (Clarity AD): NCT03887455. ClinicalTrials.gov https://clinicaltrials.gov/ct2/show/NCT03887455 (2022).

41. Chen, Z. et al. Exploring the feasibility of using real-world data from a large clinical data research network to simulate clinical trials of Alzheimer’s disease. Npj Digit. Med. 4, 84 (2021).

42. Eisai Inc. Comparison of 23 mg Donepezil Sustained Release (SR) to 10 mg Donepezil Immediate Release (IR) in Patients With Moderate to Severe Alzheimer’s Disease: NCT00478205. ClinicalTrials.gov https://www.clinicaltrials.gov/ct2/show/study/NCT00478205 (2014).

43. Jutten, R. J. et al. Finding Treatment Effects in Alzheimer Trials in the Face of Disease Progression Heterogeneity. Neurology 96, e2673–e2684 (2021).

44. Folstein, M. F., Folstein, S. E. & McHugh, P. R. “Mini-mental state”. J. Psychiatr. Res. 12, 189–198 (1975).

45. Cedarbaum, J. M. et al. Rationale for use of the Clinical Dementia Rating Sum of Boxes as a primary outcome measure for Alzheimer’s disease clinical trials. Alzheimers Dement. 9, (2013).

46. van Dyck, C. H. et al. Lecanemab in Early Alzheimer’s Disease. N Engl J Med 13 (2022).

47. The Alzheimer’s Disease Neuroimaging Initiative (ADNI). ADNI1 Procedures Manual. The Alzheimer’s Disease Neuroimaging Initiative http://adni.loni.usc.edu/wp-content/uploads/2010/09/ADNI_GeneralProceduresManual.pdf (2004).

48. The Alzheimer’s Disease Neuroimaging Initiative (ADNI). ADNI2 Procedures Manual. The Alzheimer’s Disease Neuroimaging Initiative https://adni.loni.usc.edu/wp-content/uploads/2008/07/adni2-procedures-manual.pdf (2011).

49. The Alzheimer’s Disease Neuroimaging Initiative (ADNI). ADNI GO Procedures Manual. The Alzheimer’s Disease Neuroimaging Initiative https://adni.loni.usc.edu/wp-content/uploads/2008/07/ADNI_GO_Procedures_Manual_06102011.pdf (2009).

50. The Alzheimer’s Disease Neuroimaging Initiative (ADNI). ADNI3 Procedures Manual. The Alzheimer’s Disease Neuroimaging Initiative https://clinicaltrials.gov/ct2/show/NCT02854033 (2016).

51. Bucci, M., Chiotis, K., Nordberg, A., & for the Alzheimer’s Disease Neuroimaging Initiative. Alzheimer’s disease profiled by fluid and imaging markers: tau PET best predicts cognitive decline. Mol. Psychiatry 26, 5888–5898 (2021).

52. Villeneuve, S. et al. Existing Pittsburgh Compound-B positron emission tomography thresholds are too high: statistical and pathological evaluation. Brain 138, 2020–2033 (2010).

53. Shaw, L. M. & Trojanowski, J. Q. Biomarker Core report: Year1 ADNI3, Roche Elecsys immunoassay analyses of ADNI1/GO/2 CSF samples. Alzheimer’s Association https://www.alz.org/media/documents/ww-adni-may-2017-biomarker-core-shaw.pdf.

54. Bates, D., Mächler, M., Bolker, B. & Walker, S. Fitting Linear Mixed-Effects Models Using lme4. J. Stat. Softw. 67, (2015).

55. Farrer, L. A. Effects of Age, Sex, and Ethnicity on the Association Between Apolipoprotein E Genotype and Alzheimer Disease: A Meta-analysis. JAMA 278, 1349 (1997).

56. O’Bryant, S. E. Staging Dementia Using Clinical Dementia Rating Scale Sum of Boxes Scores: A Texas Alzheimer’s Research Consortium Study. Arch. Neurol. 65, 1091 (2008).

57. Lüdecke, D. ggeffects: Tidy data frames of marginal effects from regression models. J. Open Source Softw. 3, 772 (2018).

58. Nakagawa, S. & Schielzeth, H. A general and simple method for obtaining R2 from generalized linear mixed-effects models. Methods Ecol. Evol. 4, 133–142 (2013).

59. Nakagawa, S., Johnson, P. C. D. & Schielzeth, H. The coefficient of determination R2 and intra-class correlation coefficient from generalized linear mixed-effects models revisited and expanded. J. R. Soc. Interface 14, 20170213 (2017).

60. Gelman, A. & Su, Y.-S. arm: Data analysis using regression and Multilevel/Hierarchical models. https://CRAN.R-project.org/package=arm (2022).

61. Andrews, J. S. et al. Disease severity and minimal clinically important differences in clinical outcome assessments for Alzheimer’s disease clinical trials. Alzheimers Dement. Transl. Res. Clin. Interv. 5, 354–363 (2019).

62. O’Bryant, S. E. et al. Validation of the New Interpretive Guidelines for the Clinical Dementia Rating Scale Sum of Boxes Score in the National Alzheimer’s Coordinating Center Database. Arch. Neurol. 67, (2010).

63. Efron, B. & Tibshirani, R. An introduction to the bootstrap. (Chapman & Hall, 1993).

64. Gelman, A. & Hill, J. Data analysis using regression and Multilevel/Hierarchical models. (Cambridge University Press, 2006). doi:10.1017/CBO9780511790942.

65. Baker, D. H. et al. Power contours: Optimising sample size and precision in experimental psychology and human neuroscience. Psychol. Methods 26, 295–314 (2021).

66. Bycroft, C. et al. The UK Biobank resource with deep phenotyping and genomic data. Nature 562, 203–209 (2018).

67. Shishegar, R. et al. Using imputation to provide harmonized longitudinal measures of cognition across AIBL and ADNI. Sci. Rep. 11, 23788 (2021).

68. Nagin, D. S. Group-Based Trajectory Modeling: An Overview. Ann. Nutr. Metab. 65, 205–210 (2014).

69. Donohue, M. C. & Aisen, P. S. Mixed model of repeated measures versus slope models in Alzheimer’s disease clinical trials. J. Nutr. Health Aging 16, 360–364 (2012).

